# The efficiency of dynamic regional lockdown approach in controlling the COVID-19 epidemic. Insights from the agent-based epidemiological model for Poland

**DOI:** 10.1101/2021.09.06.21263031

**Authors:** Jakub Zieliński, Magdalena Gruziel-Słomka, Jȩdrzej M. Nowosielski, Rafał P. Bartczuk, Karol Niedzielewski, Marcin Semeniuk, Łukasz Górski, Jan Kisielewski, Antoni Moszyński, Maciej Radwan, Artur Kaczorek, Franciszek Rakowski

## Abstract

In this work properties of the dynamic regional lockdown approach to suppress the COVID-19 epidemic spread in Poland were investigated. In particular, an agent based model was used with the aim to indicate an optimal lockdown strategy, defined here as the one which minimizes mean lockdown time over regional unit provided health service is not overwhelmed. With this approach the lockdown extent was also considered by varying restrictions between complete regional school closure and/or significant social distancing in semi-public spaces. In result, a cooperative effect was discovered in the case when closure of schools was accompanied by severe restrictions of social contacts in semi-public spaces. Moreover, the regional lockdown approach implemented here on the level of counties (units of population around 100k) proofed to be successful, that is allowed to identify optimal entrance and release thresholds for lockdown. The authors believe that until significant portion of population is vaccinated such a strategy might be applied.

## Introduction

The COVID-19 epidemic, which probably started at the turn of 2019 and 2020, unfortunately does not end. Although there are a few approved vaccines, vaccine supply remains an unresolved challenge. Moreover, some people refuse to be vaccinated for various reasons. Due to the high infectivity of the SARS-CoV-2 virus and a large percentage of people with asymptomatic course of illness (and therefore difficult to detect), the effective suppression of the disease requires use of drastic solutions, such as lockdown or common quarantines. The consequences of these actions are dramatic for the economy, and it seems that many countries will not be able to re-apply full lockdown due to the economic consequences.

It is therefore essential to look for the solutions that can help suppress the epidemic while reducing negative social effects. It seems that most countries will have to implement some form of smart lockdowns - they must be strong enough to prevent a sharp increase in number of infections, and weak enough not to cause a significant increase of unemployment. One of the proposed solutions is a regional dynamic lockdown strategy. In essence, it comes down to the application of strong restrictions in regions particularly strongly affected by the pandemic. After the number of infected decreases, restrictions should be relaxed in a given region.

However, managing a regional dynamic lockdown strategy is difficult. It requires the adoption of closing and opening criteria. It seems advisable to avoid both temporal oscillations (alternating switching on and off of lockdowns), and excessive mixing of closed areas with unrestricted ones — a checkerboard pattern. In order to understand inner workings of dynamic regional restrictions we use a massive agent based model (ABM) initially developed for the Polish population^1^ and equipped with a number of extensions tailored for specific characteristics of COVID-19. The model not only allows to predict the total number of cases, but also gives the information on the spatial and age distributions of infections. Model also tries to approximate the contact matrix (age of the infected person according to the age of the person who bans) or the circumstances of the infection (school, home, public transport, etc.). For the present study, we have initially tested the effectiveness of dynamic regional restricting social activity strategies with just school closures. Schools closure is a commonly used strategy to control the COVID-19 epidemic. At the peak of global schools lockdowns caused by COVID-19 (2 April 2020), 172 nations had enacted full closures or partial dismissals, affecting nearly 1.5 billion children^2^. This type of intervention is relatively simple to manage. Most often, the closure of schools depends on an administrative decision and, to a relatively small extent, depends on the social willingness to comply with it. Moreover, the current comparisons of strategies applied in the fight against COVID-19 showed a high position of school closures in the ranking of the effectiveness of non-pharmaceutical measures^3^.

In an epidemic situation, in the absence of empirical data, epidemiological models can help to evaluate the impact of interventions on transmission, and can be used as important tools to identify effective policies for appropriate measures to enact^4–8^. Based on model calculations, closure of schools was considered an effective strategy to contain epidemic of influenza^9–13^ and measles^14^, because of high contact rates among children and adolescents as well as high susceptibility to these viruses. The results of agent-based modeling of the impact of school closings on the COVID-19 epidemic in Ontario, Canada, showed that school closings may have a limited effect in reducing the burden of the disease without other measures to disrupt the transmission chain (as community-based voluntary self-isolation)^15^. Other works also confirmed the importance of introducing other measures alongside school closure, for example, related to the reduction of contacts in the workplace^5,6^.

Similarly, our results for Poland show that dynamic regional school closing alone cannot be both short in time and sufficiently effective to prevent the overload of health service. Therefore, we also consider regional and dynamic restriction on social activity in semi-public spaces (the “street” context in our model, apart from streets it covers also public and commercial services, shops etc., the “street” context does not cover schools and nurseries). Although the latter appears to be even less effective than school closing alone, we find a cooperative effect when the two contexts are locked. The effect is significant enough to make the strategy of regional dynamic restrictions imposed both on semi-public spaces and schools an attractive approach for the state authorities.

The above result is particularly important in the face of negative consequences brought by a long-term distance learning period. Not only educational, and emotional damages among children should be mentioned here, but also the secondary economic losses caused by the need for child care.

## Aim of the study

The aim of this study is twofold. Firstly, to test whether and to what extent the dynamic, regional lockdown approach is capable of suppressing the COVID-19 epidemic spread in Poland. This part of the study is done by performing number of agent-based simulations with various lockdown entrance, and release thresholds. The thresholds are derived from the number of infections, and imposed on the level of counties. Also, each series of simulations is computed for three levels of possible lockdown stringency. Secondly, provided the approach turns out successful, further analysis of simulation results aims at determining the optimal strategy of regional lockdowns. Namely, the goal is to minimize the lockdown time (averaged over counties) with a constraint to the maximum allowed occupancy of ICUs (intensive care units). For this purpose, an arbitrarily chosen critical value is assumed, corresponding to 90% of Polish public health service capacity. Since estimated ICU capacity in Poland is around 3000 (about 8 per 100 000 inhabitants), the critical value is set at 7.2 per 10^5^ inhabitants.

## Results

The following sections aim at describing the impact of school closures accompanied by restricted public places („street”) contacts on the epidemic outcome. The epidemic simulation was based on actual data until August 30, 2020. Based on data from September 2020, when schools were open in Poland, the model parameters controlling the probability of infection in schools were estimated. The beginning of the school year in Poland takes place on September 1, and from that day on, the strategies of dynamic school lockdowns (excluding kindergartens) were implemented with a county resolution. A county (*pol. powiat*) is an intermediate administrative unit in Poland and data on the development of the epidemic are collected at the county level.

There are two main parameters in the dynamic regional lockdown approach (DRLA) described in this work - the entrance and the release lockdown thresholds. These thresholds are given as the numbers of detected infected agents within two weeks period per 10^4^ agents in a county. We consider several entrance and release threshold values: 2, 4, 6, 8, 10, 12, 18, and 24 under the condition that the release threshold is less or equal than the entrance one. New cases were counted in a two-week window to avoid simple statistical fluctuations and weekly oscillations in reporting data related to the weekends. The threshold criteria were checked daily. The simulations were conducted for one school year but analyzed in detail for the first semester only.

For a reference, three additional simulations were run with countrywide restrictions being applied throughout the whole simulation: only school closure applied, only restricted street contacts applied, and both applied together.

To assess the impact of the applied restrictions, and to understand the epidemic scenarios themselves, several typical indices will be used, see Figure 1. Crucial parameters for our purpose are those that directly describe the burden of epidemic either on health system service - e.g. the interval when majority of available ICU beds are occupied or economy - e.g. the length of the lockdown interval. An attack rate does not seem to be an essential criterion for decision making on lockdown introduction, taking into account that majority of cases are asymptomatic or have a mild course of infection. However, it carries the information on the level of immunity in the society, assuming the recovered agents become immune.

**Figure 1.**
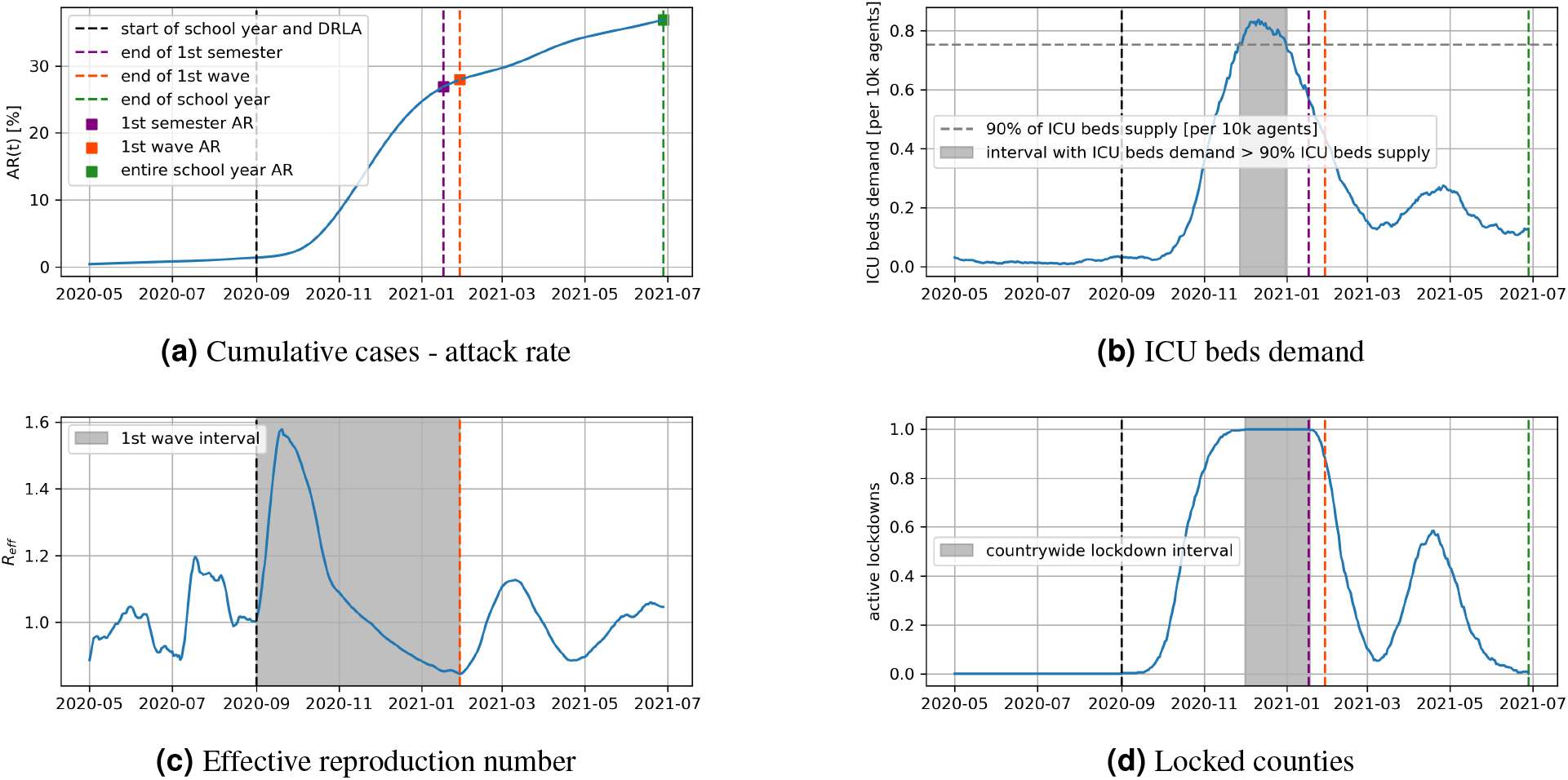
Example results - from simulation with entrance and release threshold values equal 12 and 10, respectively - describing the indices used in this paper: the attack rate during the first semester, the first wave, or the entire school year, Figure 1a; the interval over which the demand for ICU beds exceeded 90% of the supply, Figure 1b; the interval of the first wave defined via *R*_*e f f*_, Figure 1c; the interval of a full - countrywide - lockdown, Figure 1d. The vertical, dashed lines mark the start (black, dashed) or the ends of certain intervals described in the legend of Figure 1a

### The impact of lockdown entrance threshold

In this section, we consider the effect of lockdown entrance threshold (LET) on ICU overload time, and attack rate.

The attack rate till Dec., 3rd (see Figure 2a) strongly depends on LET for simulations with school closure applied (diamonds) or with school closure accompanied by restriction imposed on public spaces (squares). On the other hand, it weakly depends on LET for simulations with public spaces restricted (circles). One can observe almost 4-fold increase in the attack rate between simulations with LET 2 and 24 and both contexts restricted, and about 2.5-fold increase in simulations with only schools closed.

**Figure 2.**
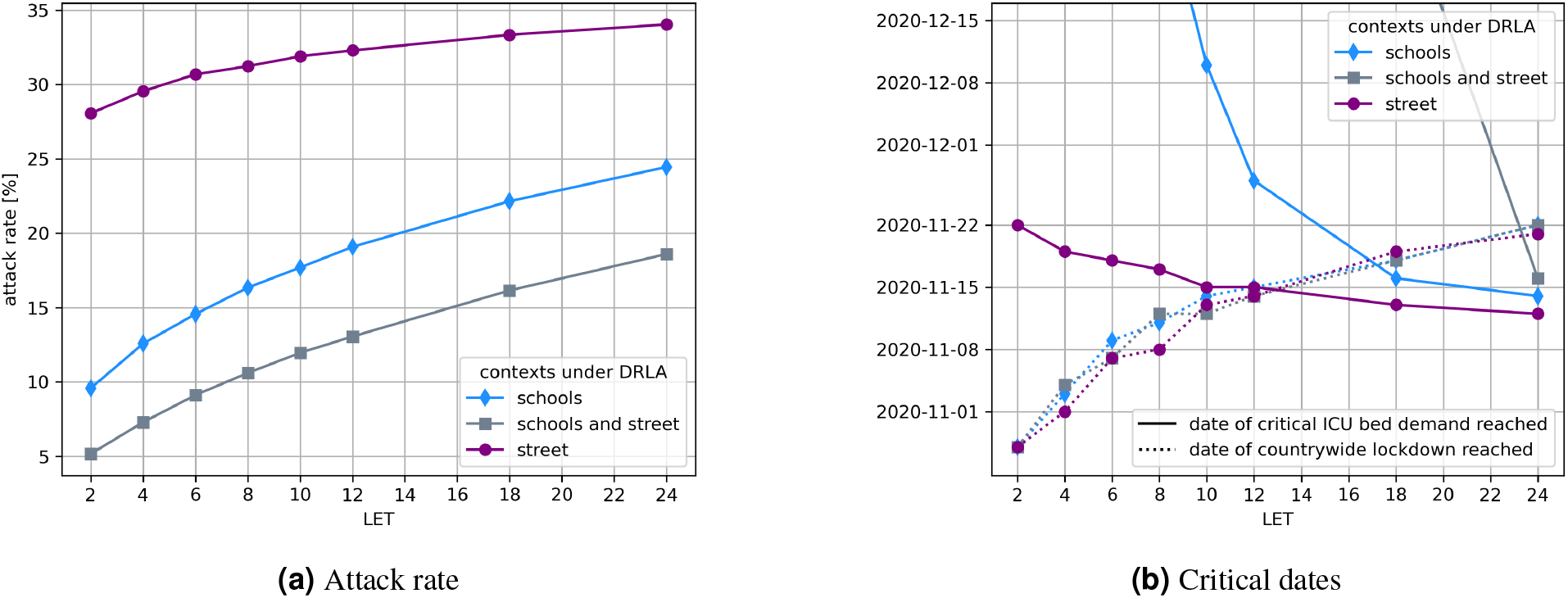
The lockdown entrance threshold dependent attack rate till Dec., 3rd, Figure 2a, and critical dates, Figure 2b. Both figures present the results for three lockdown definitions (street restricted - purple circles and lines, schools closed - blue diamonds and lines, both contexts restricted - grey squares and lines). The critical dates in Figure 2b are represented here by the day when critical ICU bed demand was reached (solid lines), and the day when countrywide lockdown was reached (dotted lines).

Clearly, the higher the lockdown entrance threshold value, the later lockdown is reached. This applies to both individual counties, and countrywide lockdown - the state in which the entry threshold has been exceeded in all counties. What may seem a bit surprising, the difference in the time of reaching the national lockdown for LET = 4 and LET = 24 is only 3 weeks, regardless of the strength of the lockdown, see Figure 2b. This is a consequence of the exponential nature of the epidemic and its suppression by regional lockdowns. However, for a very high entry threshold, national lockdown may occur later than the ICU overcapacity. This would lead to mass deaths due to lack of available care. Thus, the introduction of a strong national lockdown would be almost certain, regardless of earlier assumptions made by the government.

As one can see, closing of schools reduces the number of infections much more than closing of the street context (public places). This is due to the large number of households in Poland where three generations live together. This means that children who are infected at school and often pass infection asymptomatically, infect a significant number of household members. Moreover, classes in Poland are slightly overcrowded - e.g. in large cities there are about 25 children in the class. On the other hand, in Poland, contrary to the countries of southern Europe, regular visits to restaurants, cafes, etc. are relatively unpopular. It must be noted, however, that the impact of the lockdown imposed on public spaces is much stronger when it accompanies school closure. The relationship between the effects of school closure and restrictions imposed on public spaces is thus country dependent. However, the relationship between attack rate and LET should be universal. The higher the LET, the higher the attack rate. Furthermore the stronger the lockdown is, the more attack rate depends on the LET.

### Consecutive waves. Effective R and Lockdown Release Threshold

As a result of lockdown and achieved herd immunity, number of infections is reduced. The timing of the lockdown completion depends on lockdown release threshold (LRT). Of course, release of lockdown could lead to a new wave of infections. When multiple epidemic waves occur, it is necessary to define a boundary between them. In this paper, as the beginning of a new wave, we assume the moment when effective reproduction number *R*_*e f f*_ starts to rise. Therefore, in order to observe the occurrence of successive waves over time, it is convenient to look at the graph showing the variability of *R*_*e f f*_.

As one may notice (see Figure 3), both LET and LRT strongly influence the times of the beginning and end of successive waves of epidemics. A stronger lockdown means a greater risk of next epidemic waves coming soon. Someone might consider this a disadvantage of strong lockdown or an advantage of a weak one. However, this is an apparent benefit resulting from the ineffectiveness of a weak lockdown. Weak lockdown, as well as its absence, lead to a high *R*_*e f f*_ value, long first wave and a large number of infections in short time, and thus unacceptably many deaths.

**Figure 3.**
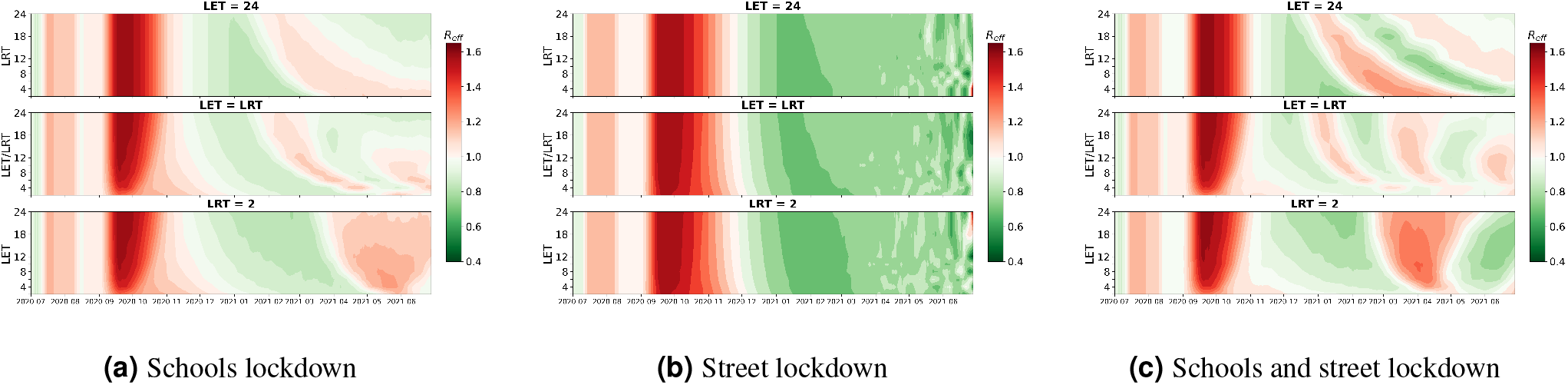
Change of effective reproduction number, *R*_*e f f*_, in time for various combinations of LET and LRT thresholds. Panels in each figure correspond to: LRT dependent *R*_*e f f*_ (*t*) for LET=24 (top), LET dependent *R*_*e f f*_ (*t*) for LRT=2 (bottom), and *R*_*e f f*_ (*t*) for LET=LRT (middle).

As one can see, in the case of a strong lockdown, especially for a high LET value, the occurrence times of subsequent epidemic waves strongly depend on the LRT. This means that the times when the first wave ends, and the next waves occur depend on government decisions. On the other hand, governments have no influence on the starting time of the first wave. Moreover, as shown in the previous section, the choice of particular LET can shift the beginning of the first lockdown just by few weeks.

### Searching for an optimal lockdown strategy

In many countries, completely different lockdown strategies have been chosen, which means that choosing the optimal one is not easy. On the one hand, we all know that too long lockdowns are socially unacceptable. On the other hand, the capacity of health systems must not be exceeded. These needs seem contradictory.

In order to find the optimal lockdown strategy, we ran the model with different LET and LRT threshold values. The simulation results are presented on Fig. 4. As one can see, in the case of a weak lockdown (just closing public places or schools only, see Fig. 4a, 4b) there is a strong correlation between the total lockdown time (background shades) and the time when ICUs are overcrowded (quares color). Both ICU overcrowding time duration of lockdowns strongly depend on LET and quite weakly on LRT. Therefore, the application of weak lockdowns does not allow avoiding the dilemma: economic losses vs a large number of deaths.

**Figure 4.**
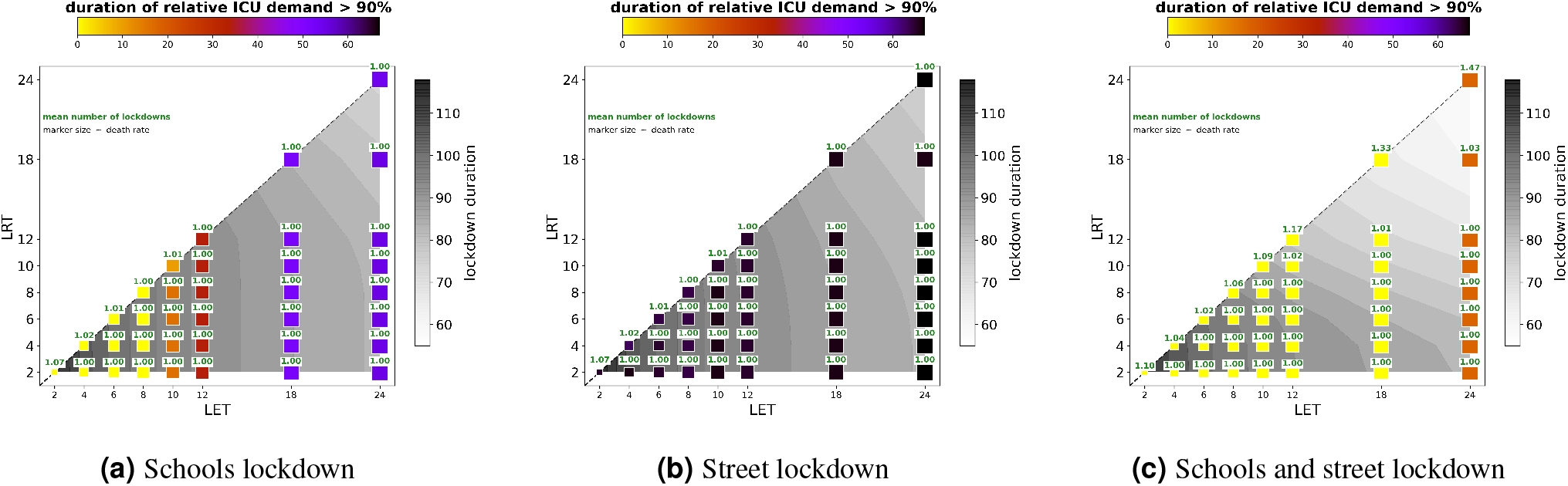
Various indices dependent on LET and LRT values: grey scale contours correspond to the mean (over counties) lockdown duration in days, marker colors (horizontal colorbar) correspond to the duration of period when relative ICU bed demand exceeded 90% of ICU bed supply (in days). Numbers above markers (green font) give the mean (over counties) number of lockdown events, and finally marker size roughly corresponds to death rate. All indices where calculated for 1st semester interval.

Fig. 4c shows the effect of applying a strong lockdown. Obviously, the lockdown time is shorter, as is the ICU overload time. Of course, this is not surprising. However, in a strong lockdown, increasing the LRT significantly shortens the lockdown time, leaving the ICU overcapacity time almost unchanged. This is a very different situation from a case with a weak lockdown imposed. For example, for LET = 18, increasing the LRT from 2 to 18 reduces the total lockdown duration by approximately two weeks. The most serious consequence is increasing the average number of lockdowns in the county from 1 to 1.33. This means that some poviats will have to apply a repeated, short lockdown. The cost seems low.

### More detailed analysis of lockdowns

An important feature of the proposed scheme is the regionalization of lockdowns, as it allow to prevent the introduction of civil restrictions where and when they are not not necessary. Therefore, the transition of regional lockdowns into a countrywide lockdown, i.e. the situation of closing all counties, is in a sense a failure of the proposed strategy. The Fig. 5 shows the time of the countrywide lockdown in relation to the average time of closing individual poviats. As one can see, only in the case of a strong lockdown and high thresholds, countrywide lockdown does not occur (Fig. 5c).

**Figure 5.**
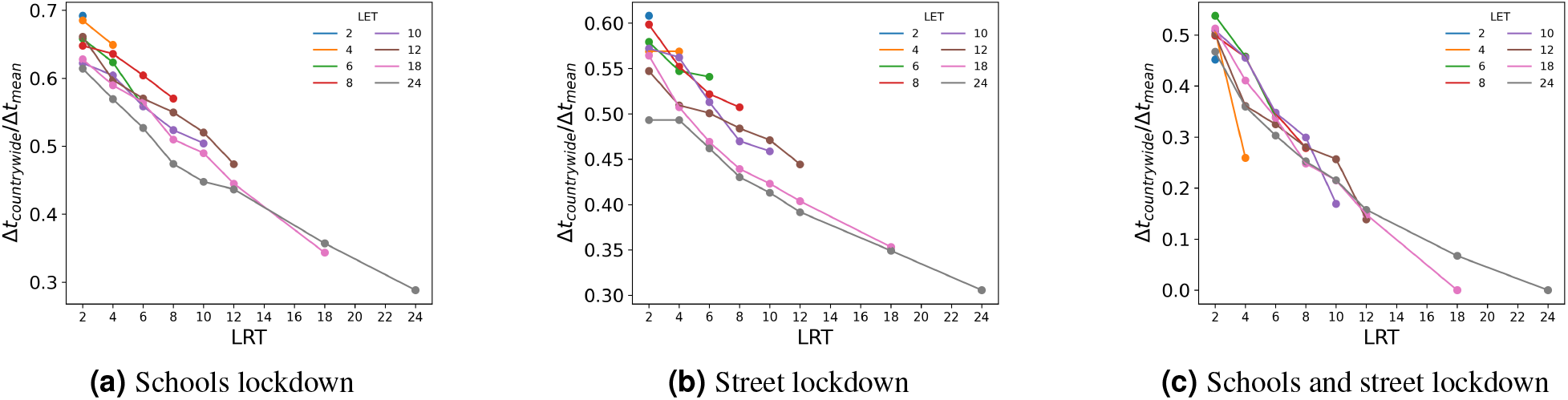
The ratio of countrywide lockdown interval to mean (over counties) lockdown interval. The high value of this coefficient is associated with the failure of the regional lockdown strategy.

Finally, we analyzed the sensitivity of the proposed strategy to the threshold values. The green bar in Fig. 6 shows the relative ICU demand as a function of time for different thresholds. A wide bar at a given moment in time means that the number of people needing the ventilator is highly sensitive to the LRT and LET values. This is important as the threshold values for critical ICU demand can vary significantly between countries. As can be seen, in Fig. 6c, in the case of a strong regional lockdown (both schools and public places - streets are closed), it is possible to maintain the number of occupied respirators at an acceptable level, regardless of the adopted, reasonable LET and LRT values. On the other hand, a removal of a strong, countrywide lockdown threatens with the next wave of an epidemic. In this case/In such a case, a countrywide moderate lockdown (i.e. in this work, school closure only) appears to be a reasonable alternative.

**Figure 6.**
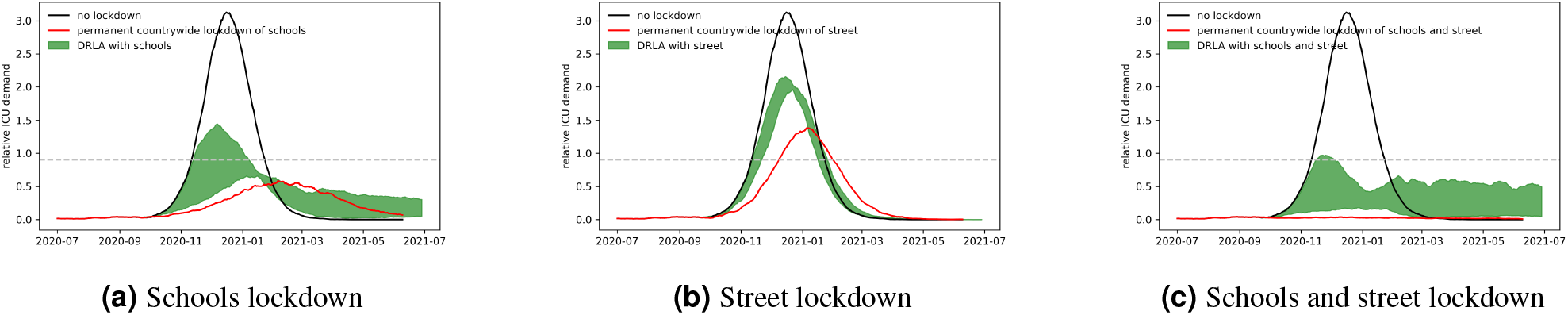
Comparison of time dependent relative ICU demand for all scenarios with DRLA on selected contexts (shaded green region), with scenario of countrywide permanent lockdown of these contexts (red line), and with scenario with no lockdown applied (black line). Shaded region marks the bounds between minimum and maximum values obtained in all DRLA scenarios (with selected contexts restricted). Dashed horizontal line marks the critical ICU bed demand.

## Discussion and summary

The experience of many Asian countries and, among others, New Zealand, which managed to suppress the SARS-Cov-2 virus epidemic, show that the most effective method of fighting this virus is a very strong lockdown or contact tracing and mass testing. Unfortunately, such a strategy has not been implemented in most European and American countries. Usually, there are cultural and legal reasons that impede the introduction of absolute lockdowns and tracking citizens’ contacts. Testing was limited by economic factors and the lack of availability of a large number of tests. The huge number of infections in India and Brazil shows the scale of the vaccine shortage. Therefore, for many countries, a different, realistic strategy is needed.

For economic and social reasons, governments are defending themselves against ordering strong and long lockdowns. It is obvious to most people that the more costly (economically and socially) lockdown is, the more effective it is. So a dilemma arises whether to sacrifice people’s lives or the economy and, among others, the psyche of children. In this study we have shown that a possible strong, regional and short lockdown allows governments to avoid the above dilemma. Strong lockdown, even if it does not eradicate the epidemic, may suppress it enough to allow for the lockdown release. Of course, a strong lockdown itself has many unpleasant consequences. Therefore, it is crucial to avoid introducing lockdowns when and where they are not necessary. Therefore, they should be introduced and lifted at the appropriate moment and only in parts of the country where it is necessary. In our opinion, the strategy is the optimal method of epidemic management until the population is vaccinated.

## Methods

This section briefly describes the agent-based model used in this paper.

### Model description

The results presented in this work were obtained with the usage of an agent based model. Initially, the model was developed to explain influenza epidemic in Poland^1^. Similarly to influenza, Covid-19 is an infectious airborne disease. In order to adapt the model for simulation of the Covid-19 epidemic spread a number of modifications, and new features were introduced.

Epidemic spread takes place within the virtual society of Poland^16^. The virtual society consists of 36 million geo-referenced software agents. Agents meet in various contexts where they can contract the disease. Each agent is assigned to exactly one geo-referenced household context. A workplace, a preschool, a school, a university, a large university, a street, and a travel are the other context categories present in the model. Contexts are characterised by contacting rates (CRs), which are model parameters. One can therefore model the imposing and lifting of governmental restrictions by changing the CRs. Every agent attends contexts he/she belongs to, and after a full day the probability of infection for each susceptible agent is calculated. The probability, expressed simply as:

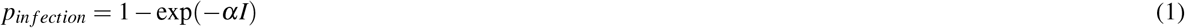

depends on several time dependent parameters either occasionally changing, such as coefficient *f*, and contacting rates (*w*_*c*_) of contexts, or changing every timestep, such as context infectivities, i.e., proportion of infectious agents present in the given context. It also depends on constant parameters, i.e. transmission coefficient *α*. Transmission coefficient *α* is a parameter of COVID-19 itself and coefficient *f* describes fraction of infectious symptomatic agents who do not self-isolate despite the fact that they feel unwell.

The first important extension of the model is the introduction of asymptomatic infected agents. The motivation is to take into account COVID-19 cases which do not exhibit any (obvious) symptoms of the disease for the whole course of the infection. We assume that infectiousness of an asymptomatic agent is multiplied by parameter *a* (i.e. *a* = 0.1) with respect to a symptomatic agent. The proportion of asymptomatic and symptomatic agents varies across agents age category.

With the above, the context infectivities become:

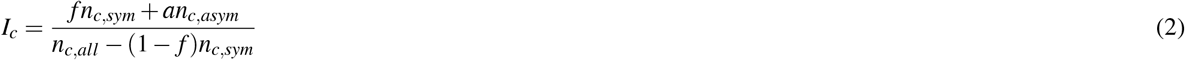

where *c* stands for a household, a workplace, a preschool, a school, a university, a large university, a street, or a travel. For a household, the *f* parameter is assumed to be 1. The total infectivity, *I*, that enters the Eq. 1 is just:

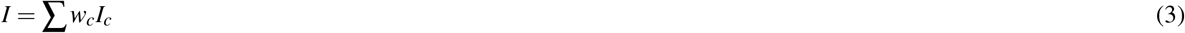

where *w*_*c*_ are the contacting rates within a given context, and the sum goes over all the contexts a given agent visited/attended during a given day.

Once infected, an agent may follow a number of possible paths over health states, see Fig. 7. Upon contagion an agent becomes latent. Next, the infected agent might enter either symptomatic or asymptomatic course of infection. For the symptomatic case, three paths of various severity are possible: the infection with light symptoms without the necessity of hospitalisation, the infection with severe symptoms calling for the hospitalisation, and finally the infection course that leads to admittance to intensive care unit (ICU).

**Figure 7.**
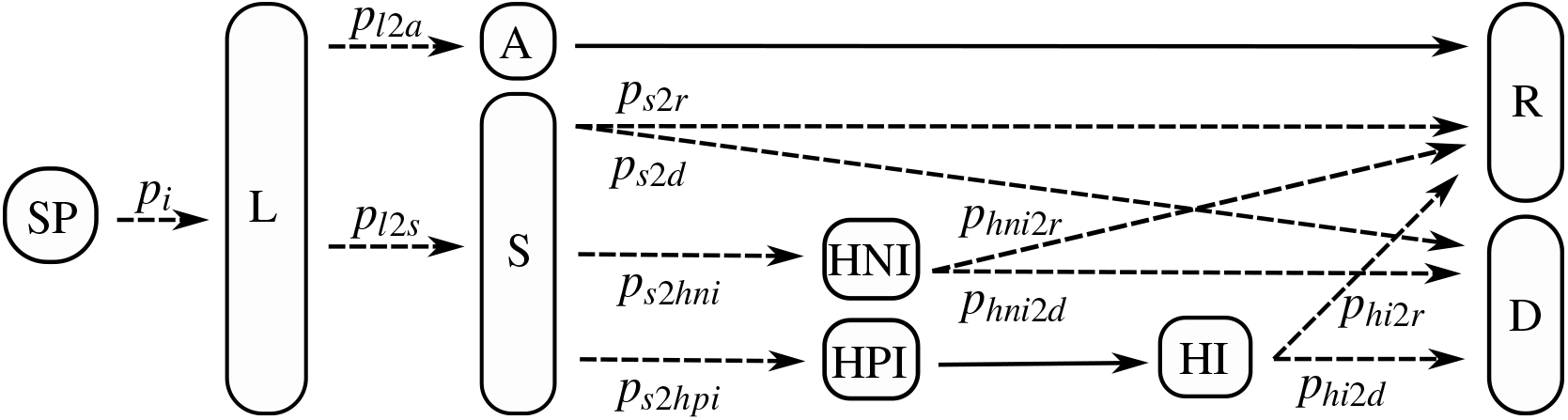
The possible paths through the course/states of illness. The state abbreviations stand for: SP - susceptible, A - asymptomatic, S - symptomatic, HPI - hospitalized, pre ICU, HI - hospitalized, at ICU, HNI - hospitalized, not at ICU, D - dead, R - recovered. There are two certain transitions (with transition probability equal 1; marked with solid arrows) regardless of the agent’s age. The general probability of infection, *p*_*i*_ (the probability of the transition from susceptible to latent state; dashed arrow) depends on various parameters described in the text, and the probabilities of the rest of the transitions (also dashed arrows) are only age dependent, and are collected in Table 1.

COVID-19 is known for strong correlation between disease severity and age, which leads to the introduction of another important model extension where the probabilities of the transitions between the health states (e.g. the probability of asymptomatic infection course) depend on the agent age. These transition probabilities are gathered in Tab. 1.

**Table 1.**

| age range | $p_{l2a}$ | $p_{l2s}$ | $p_{s2hni}$ | $p_{s2hpi}$ | $p_{hni2d}$ | $p_{hi2d}$ | $p_{s2d}$ | $p_{hni2r}$ | $p_{hi2r}$ | $p_{s2r}$ |
| --- | --- | --- | --- | --- | --- | --- | --- | --- | --- | --- |
| 0 - 10 | 0.95 | 0.05 | 0.01 | 0.001 | 0.08 | 0.75 | 0.001 | 0.92 | 0.25 | 0.988 |
| 10 - 20 | 0.95 | 0.05 | 0.01 | 0.001 | 0.08 | 0.75 | 0.001 | 0.92 | 0.25 | 0.988 |
| 20 - 30 | 0.95 | 0.05 | 0.012 | 0.002 | 0.08 | 0.75 | 0.001 | 0.92 | 0.25 | 0.985 |
| 30 - 40 | 0.9 | 0.1 | 0.018 | 0.003 | 0.08 | 0.75 | 0.002 | 0.92 | 0.25 | 0.977 |
| 40 - 50 | 0.9 | 0.1 | 0.035 | 0.005 | 0.08 | 0.75 | 0.002 | 0.92 | 0.25 | 0.958 |
| 50 - 60 | 0.8 | 0.2 | 0.07 | 0.01 | 0.08 | 0.75 | 0.005 | 0.92 | 0.25 | 0.915 |
| 60 - 70 | 0.77 | 0.23 | 0.2 | 0.05 | 0.08 | 0.75 | 0.02 | 0.92 | 0.25 | 0.73 |
| 70 - | 0.52 | 0.48 | 0.25 | 0.1 | 0.08 | 0.75 | 0.03 | 0.92 | 0.25 | 0.62 |

There is one more important issue related to the transitions between health states, namely the time periods of the states. One should note here, that the splitting into states is already arbitrary. Based on available data, the time periods of the infection states (e.g. length of latent state - the time before the onset of symptoms/infectivity - or the hospitalization time) are described by time distributions rather than fixed values. Even these distributions probably vary with the age of infected person. However, for the simplicity, this model considers only fixed, age-independent values of health state periods, see Tab. 2. The coefficients collected in Tables 1 and 2 were defined using literature data^17–19^ and information collected by the National Institute of Public Health based on data from Polish hospitals.

**Table 2.**

| L | A | S | HNI | HPI | HI | D | R |
| --- | --- | --- | --- | --- | --- | --- | --- |
| 4 | 7 | 5 | 10 | 13 | 7 | ∞ | ∞ |

Introduction of quarantine status variable is another model extension. It allows to simulate administrative order of isolation imposed on agents with confirmed infection as well as quarantine imposed on other agents (e.g. agents who were in contact with an infected agent or agents who came back from abroad). A quarantine status variable is independent of the health state variable. It represents the isolation and quarantine enforced by the state authorities in contrary to coefficient *f* which is related to the common sense self-isolation of the symptomatic infected agents. Given proportion of infected symptomatic agents are placed under quarantine. This reflects the fact that majority of cases are not detected by authorities. If a given agent is placed under quarantine his/her household members are as well. Only a fraction *q* of infected agents are placed under quarantine. This reflects the fact that many cases are not detected by authorities.

### Model calibration

The model calibration consists in determining the model parameter values in order to obtain consistency with the real COVID-19 epidemic spread which occurred in Poland until the end of school summer holidays, i.e., until 31st of August 2020. The main focus of the paper is to analyse hypothetical dynamic school closures starting from 1st of September 2020. The considered dynamic school closure policy is not directly comparable to the real epidemic course in Poland after the start of the school year. While we are still working on formal model verification procedures, it should be noted that it was already validated against real-world data^20^, in Polish-German hub^21^.

In the calibration process one can distinguish two categories of model parameters. Transmission coefficient *α* and coefficient *f* were optimized using Bayesian optimization.^22^ Finally, contacting rates in contexts were determined by expert informed trial and error method.

The initial contacting rates [CR] were adopted from Rakowski et al.^1^. CR changes during the outbreak and subsequent restrictions were implemented through multipliers (*µ*_*c*_ ≥ 0, where *c* represents context). Introduction of the multipliers changes the Eq. 3 into 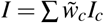, with 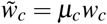. In order to model changes in the CR for a particular context, we utilized the trial and error method. For preschools (kindergartens, *µ*_*KG*_), schools (*µ*_*SCH*_), universities (*µ*_*EU*_), and large universities contexts (*µ*_*LEU*_), these multipliers were estimated based on the proportion of people attending these places after introducing a given restriction. The multipliers of long-distance traveling-related CRs (number of co-travelers - *µ*_*COT*_, probability to travel - *µ*_*TRAV*_) were based on the content of government decisions regarding the use of public transport, determining the maximum number of seats taken. The multipliers of the *f* parameter, which quantifies common-sense self-isolation and self-isolation resulting from restrictions, were based on the assumption of increasing COVID-19 awareness in society and changes in penalties for moving COVID-19-positive individuals in the public space. CR multipliers for the households (*µ*_*HH*_), workplaces (*µ*_*WP*_), and street contexts (*µ*_*STR*_) were adjusted with an educated guess, assuming that household CRs partly replaced those at workplaces, education, or streets.

In the trial and error experiments, the minimum and maximum values of the multipliers related to the introduction of a given restriction were first established in order to predict the course of the epidemic. Then, within the chosen range, several values of multipliers were tested to compare the results with the real-world data of identified cases from 14 days after the introduced change. Ten days is the minimum time needed to see the effect of the change in the number of confirmed cases in the model. It results from the latent time (4 days) and the time assumed for the case to be reported (6 days). Additional four days allowed to confirm changes in the trajectory. In order to determine the best set of multipliers, the Fréchet distance between the number of confirmed cases from the model and from the real-world data was minimized.

The multipliers were estimated this way until the change on September 9, when schools were already opened. (In Poland, schools were closed since March until the end of August. For the above procedure to work, a certain period when the schools were open in real life was necessary to determine CRs of the schools context).

The final values of CR multipliers are listed in Table 3.

**Table 3.**

| | $\mu_{HH}$ | $\mu_{KG}$ | $\mu_{SCH}$ | $\mu_{WP}$ | $\mu_{EU}$ | $\mu_{LEU}$ | $\mu_{COT}$ | $\mu_{STR}$ | $\mu_f$ | $\mu_{TRAV}$ |
| --- | --- | --- | --- | --- | --- | --- | --- | --- | --- | --- |
| 05.03.2020 | 1.0 | 1.0 | 1.0 | 1.0 | 1.0 | 1.0 | 1.0 | 1.0 | 1.0 | 1.0 |
| 12.03.2020 | 1.005 | 0.7525 | 0.7525 | 0.9375 | 0.0 | 0.0 | 1.0 | 0.9375 | 0.8 | 1.0 |
| 14.03.2020 | 1.02 | 0.01 | 0.01 | 0.75 | 0.0 | 0.0 | 1.0 | 0.75 | 0.2 | 1.0 |
| 24.03.2020 | 1.03 | 0.01 | 0.01 | 0.35 | 0.0 | 0.0 | 0.25 | 0.35 | 0.1 | 0.75 |
| 01.04.2020 | 1.04 | 0.01 | 0.01 | 0.2 | 0.0 | 0.0 | 0.25 | 0.2 | 0.03 | 0.5 |
| 06.04.2020 | 1.04 | 0.0 | 0.0 | 0.2 | 0.0 | 0.0 | 0.25 | 0.2 | 0.03 | 0.5 |
| 11.04.2020 | 1.0425 | 0.0 | 0.0 | 0.2 | 0.0 | 0.0 | 0.25 | 0.175 | 0.03 | 0.5 |
| 16.04.2020 | 1.05 | 0.0 | 0.0 | 0.2 | 0.0 | 0.0 | 0.25 | 0.1 | 0.03 | 0.5 |
| 20.04.2020 | 1.05 | 0.0 | 0.0 | 0.2 | 0.0 | 0.0 | 0.25 | 0.2 | 0.03 | 0.5 |
| 06.05.2020 | 1.04 | 0.35 | 0.0 | 0.2 | 0.0 | 0.0 | 0.25 | 0.2 | 0.03 | 0.5 |
| 18.05.2020 | 1.04 | 0.35 | 0.05 | 0.21 | 0.0 | 0.0 | 0.25 | 0.21 | 0.03 | 0.55 |
| 30.05.2020 | 1.0192 | 0.35 | 0.05 | 0.21 | 0.0 | 0.0 | 0.25 | 0.2205 | 0.03 | 0.55 |
| 05.06.2020 | 1.0 | 0.35 | 0.05 | 0.2 | 0.0 | 0.0 | 0.25 | 0.1 | 0.03 | 0.55 |
| 28.06.2020 | 0.9875 | 0.2875 | 0.0 | 0.2 | 0.0 | 0.0 | 0.2875 | 0.2 | 0.035 | 0.6 |
| 03.07.2020 | 0.95 | 0.1 | 0.0 | 0.2 | 0.0 | 0.0 | 0.4 | 0.5 | 0.05 | 0.75 |
| 01.08.2020 | 0.95 | 0.2 | 0.0 | 0.2 | 0.0 | 0.0 | 0.3 | 0.3 | 0.05 | 0.75 |
| 03.09.2020 | 1.0 | 0.65 | 0.65 | 0.2 | 0.0 | 0.0 | 0.3 | 0.3 | 0.05 | 0.75 |
| 30.11.2020 | 1.0 | 0.65 | 0.0 | 0.2 | 0.0 | 0.0 | 0.3 | 0.1 | 0.05 | 0.75 |

## Supporting information

Supplementary animation

## Data Availability

No data available.

## Supplementary materials

To show how the epidemic is spreading across the country, we present an example animation. It is a visualization of lock down counties for example thresholds LET = LRT = 12.

## Additional information

### Competing interests (mandatory statement)

Authors have no competing interests as defined by Nature Research, or other interests that might be perceived to influence the interpretation of the article.

## References

1. Rakowski, F., Gruziel, M., Łukasz Bieniasz-Krzywiec & Radomski, J. P. Influenza epidemic spread simulation for Poland — a large scale, individual based model study. Phys. A: Stat. Mech. its Appl. 389, 3149–3165, DOI: https://doi.org/10.1016/j.physa.2010.04.029 (2010).

2. UNESCO. Global monitoring of school closures. https://en.unesco.org/covid19/educationresponse#schoolclosures (2020). Accesed: 2021-04-14.

3. Brauner, J. M. et al. Inferring the effectiveness of government interventions against COVID-19. Science DOI: https://doi.org/10.1126/science.abd9338 (2020).

4. Aleman, D. M. et al. morPOP: a fast and granular agent-based model of COVID-19 to examine school mitigation strategies in newfoundland & labrador. In Proceedings of the 30th Annual International Conference on Computer Science and Software Engineering, CASCON’20, 266–267 (IBM Corp., 2020).

5. Germann, T. C. et al. Using an agent-based model to assess k-12 school reopenings under different COVID-19 spread scenarios – united states, school year 2020/21. medRxiv DOI: 10.1101/2020.10.09.20208876 (2020).

6. Lazebnik, T. & Bunimovich-Mendrazitsky, S. The signature features of COVID-19 pandemic in a hybrid mathematical model - implications for optimal work-school lockdown policy. medRxiv DOI: 10.1101/2020.11.02.20224584 (2020).

7. McGee, R. S., Homburger, J. R., Williams, H. E., Bergstrom, C. T. & Zhou, A. Y. Model-driven mitigation measures for reopening schools during the COVID-19 pandemic. medRxiv DOI: 10.1101/2021.01.22.21250282 (2021).

8. Li, J. & Giabbanelli, P. J. Identifying synergistic interventions to address covid-19 using a large scale agent-based model. medRxiv DOI: 10.1101/2020.12.11.20247825 (2020).

9. Fung, I. C.-H. et al. Modeling the effect of school closures in a pandemic scenario: Exploring two different contact matrices. Clin. Infect. Dis. 60, S58–S63, DOI: 10.1093/cid/civ086 (2015).

10. Milne, G. J., Kelso, J. K., Kelly, H. A., Huband, S. T. & McVernon, J. A small community model for the transmission of infectious diseases: Comparison of school closure as an intervention in individual-based models of an influenza pandemic. PLOS ONE 3, e4005, DOI: 10.1371/journal.pone.0004005 (2008).

11. Xue, Y., Kristiansen, I. S. & de Blasio, B. F. Dynamic modelling of costs and health consequences of school closure during an influenza pandemic. BMC Public Heal. 12, 962, DOI: 10.1186/1471-2458-12-962 (2012).

12. Towers, S., Geisse, K. V., Tsai, C.-C., Han, Q. & Feng, Z. The impact of school closures on pandemic influenza: Assessing potential repercussions using a seasonal SIR model. Math. Biosci. & Eng. 9, 413–430, DOI: 10.3934/mbe.2012.9.413 (2012).

13. Cauchemez, S., Valleron, A.-J., Boëlle, P.-Y., Flahault, A. & Ferguson, N. M. Estimating the impact of school closure on influenza transmission from sentinel data. Nature 452, 750–754, DOI: 10.1038/nature06732 (2008).

14. Hunter, E. & Kelleher, J. D. Using a hybrid agent-based and equation based model to test school closure policies during a measles outbreak. BMC Public Heal. 21, 499, DOI: 10.1186/s12889-021-10513-5 (2021).

15. Abdollahi, E., Haworth-Brockman, M., Keynan, Y., Langley, J. M. & Moghadas, S. M. Simulating the effect of school closure during COVID-19 outbreaks in ontario, canada. BMC Medicine 18, 230, DOI: 10.1186/s12916-020-01705-8 (2020).

16. Rakowski, F., Gruziel, M., Krych, M. & Radomski, J. P. Large scale daily contacts and mobility model — an individual-based countrywide simulation study for poland. J. Artif. Soc. Soc. Simul. 13, DOI: https://doi.org/10.18564/jasss.1529 (2010).

17. Ahrenfeldt, L. J., Otavova, M., Christensen, K. & Lindahl-Jacobsen, R. Sex and age differences in COVID-19 mortality in europe. Research Square DOI: 10.21203/rs.3.rs-61444/v1 (2020).

18. Du, R.-H. et al. Predictors of mortality for patients with COVID-19 pneumonia caused by SARS-CoV-2: a prospective cohort study. Eur. Respir. J. 55, 2000524, DOI: 10.1183/13993003.00524-2020 (2020).

19. Russell, T. W. et al. Estimating the infection and case fatality ratio for coronavirus disease (COVID-19) using age-adjusted data from the outbreak on the diamond princess cruise ship, february 2020. Eurosurveillance 25, DOI: 10.2807/1560-7917.es.2020.25.12.2000256 (2020).

20. Ramakrishna, S., Górski, Ł. & Paschke, A. A dialogue between a lawyer and computer scientist: the evaluation of knowledge transformation from legal text to computer-readable format. Appl. Artif. Intell. 30, 216–232 (2016).

21. Bracher, J. et al. A pre-registered short-term forecasting study of covid-19 in germany and poland during the second wave – publication approved by nature communications. medRxiv DOI: 10.1101/2020.12.24.20248826 (2020).

22. Shahriari, B., Swersky, K., Wang, Z., Adams, R. P. & de Freitas, N. Taking the human out of the loop: A review of bayesian optimization. Proc. IEEE 104, 148–175, DOI: 10.1109/jproc.2015.2494218 (2016).

